# Transmission potential of the novel coronavirus (COVID-19) onboard the Diamond Princess Cruises Ship, 2020

**DOI:** 10.1101/2020.02.24.20027649

**Authors:** Kenji Mizumoto, Gerardo Chowell

**Affiliations:** Graduate School of Advanced Integrated Studies in Human Survivability, Kyoto University Yoshida-Nakaadachi-cho, Sakyo-ku, Kyoto, Japan; Hakubi Center for Advanced Research, Kyoto University, Yoshidahonmachi, Sakyo-ku,Kyoto, Japan; Department of Population Health Sciences, School of Public Health, Georgia State University, Atlanta, Georgia, USA

**Author notes:** Correspondence to: K Mizumoto, Graduate School of Advanced Integrated Studies in Human Survivability, Kyoto University Yoshida-Nakaadachi-cho, Sakyo-ku, Kyoto, Japan.

## Abstract

An outbreak of COVID-19 developed aboard the Princess Cruises Ship during January-February 2020. Using mathematical modeling and time-series incidence data describing the trajectory of the outbreak among passengers and crew members, we characterize how the transmission potential varied over the course of the outbreak. Our estimate of the mean reproduction number in the confined setting reached values as high as ∼11, which is higher than mean estimates reported from community-level transmission dynamics in China and Singapore (approximate range: 1.1-7). Our findings suggest that *R*_t_ decreased substantially compared to values during the early phase after the Japanese government implemented an enhanced quarantine control. Most recent estimates of *R*_t_ reached values largely below the epidemic threshold, indicating that a secondary outbreak of the novel coronavirus was unlikely to occur aboard the Diamond Princess Ship.

## Introduction

While the novel coronavirus (Covid-19) spread rapidly throughout China for several weeks since December 2019, the virus had not taken off outside China in part due to the unprecedented social distancing measures that the Chinese government put in place. One exception is the outbreak of COVID-19 that developed aboard the Diamond Princess Ship which was detected in early February when one of its passengers, a traveler from Hong Kong, tested positive for the novel coronavirus. The number of cases in the Diamond Princess Ship quickly jumped to 454 confirmed cases by February 18, 2020. In contrast, the total number of cases in Singapore, one of the countries with the highest number of COVID-19 cases after China, was only 77 at the time [1].

Accumulating evidence indicates that the novel coronavirus can spread widely in confined settings including hospitals [2], cruise ships [3], prisons, and churches [4-5]. In Wuhan City, China, outbreaks inside health care settings led to the infection of hundreds of health professionals [2]. In Tokyo, Japan, most of the reported infections have been linked to a party inside a traditional wooden ship, called Yakatabune [6] while most of the infections in Korea have affected members of one church and one hospital.

Tracking the evolution of the transmission potential of COVID-19 in different confined settings and how it compares with that of other respiratory diseases such influenza has public health implications. When outbreaks occur in confined settings, it is useful to investigate how the effective reproduction number changes as a result of interventions strategies such as the quarantine that the Japanese government imposed on travelers and crew members aboard the Diamond Princess Ship.

In this study we sought to characterize the temporal variation in the transmission potential of the COVID-19 outbreak aboard the Princess Cruises Ship using mathematical modeling and time-series incidence data by dates of symptoms onset describing the trajectory of the outbreak among passengers and crew members.

### Epidemiological incidence cases

In Yokohama, Japan, an outbreak of COVID-19 has been unfolding on board the Princess Cruise Ship, which has been under quarantine orders since February 5, 2020, after a former passenger of the Diamond Princess Ship tested positive for the virus after disembarking in Hong Kong [3,7]. As of February 22, 2020, two days after the scheduled two-week quarantine came to an end, a total of 621 symptomatic and asymptomatic people including one quarantine officer, one nurse and one administrative officer tested positive for COVID-19 out of the 3,711 passengers and crew members on board. Laboratory tests by PCR that prioritize symptomatic or high-risk groups have been conducted.

Incidence curves of laboratory-confirmed symptomatic cases of COVID-19 among passengers and crew members of the outbreak unfolding aboard the Princess Cruises Ship are publicly available from the National Institute of Infectious Diseases, Japan (NIID) website [3]. Daily time series of symptomatic patients, from January 20, 2020 to February 18, 2020 were extracted. However, to reconstruct the trajectory of the epidemic, only 197 cases have dates of symptoms onset are available out of a total of 300 confirmed symptomatic cases.

Of the 103 symptomatic cases with missing onset dates, a total of the 79 cases are passengers while 24 cases are crew members. Moreover, out of the 79 passenger cases, 30 cases were reported from February 4, 2020 to February 6, 2020, 21 cases were reported from February 7, 2020 to February 14, 2020, and 28 cases were reported from February 14, 2020 to February 19, 2020. Of 24 crew member cases, 1 case was reported from February 4, 2020 to February 6, 2020, 15 cases were reported from February 7, 2020 to February 14, 2020, and 8 cases were reported from February 14, 2020 to February 19, 2020.

### Epidemiological modelling

We connected the daily incidence series with a discrete–time integral equation to describe the epidemic dynamics aboard the Diamond Princess Ship. Specifically, let *f*_s_ denote the probability mass function of the serial interval of COVID-19, where the serial interval is defined as the time from illness onset in the primary case to time of illness onset in the secondary case. Then *f*_s_, of length *s* days, is given by

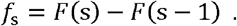

For *s*>0, *F*(s) represents the cumulative distribution function of the gamma distribution. We characterized the expected number of new incident cases E[*c*_*i,t*_] in type *i* at symptom onset week *t* as follows,

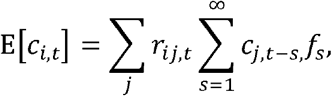

where *r*_ij_ denotes the average number of cases of type *i* infected by a single individual of type *j*. Here we assume that the incidence, *<u>c</u>*_i,t_, follows a Poisson sampling process with expected value E[*c*_i,t_].

The reproduction matrix for each type is given by

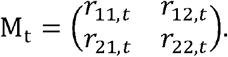

This matrix is referred to as a next-generation matrix (NGM) in a fully susceptible population [8]. Using this matrix, we derive the instantaneous time-dependent effective reproduction number, *R*_t_, for the entire transmission dynamics from the largest eigenvalue of the NGM. Under the assumption that the per-contact infection probability and the generation interval are consistent over time regardless of the type of infection, the NGM quantifies the within type and inter-type patterns of transmission [9]. The sum of the value in column *j* is the reproduction number for a specific type *j*.

Serial interval estimates of COVID-19 were derived from previous studies of COVID-19, indicating that it follows a gamma distribution with the mean and SD at 7.5 and 3.4 days, respectively, based on ref. [10]. The maximum value of the serial interval was fixed at 20 days as the cumulative probability distribution of the gamma distribution up to 20 days reaches 0.991.

We estimated model parameters and made projections using a Monte Carlo Markov Chain (MCMC) method in a Bayesian framework. Point estimates and corresponding 95% credibility intervals were drawn from the posterior probability distribution. All statistical analyses were conducted in R version 3.5.2 (R Foundation for Statistical Computing, Vienna, Austria) and the ‘rstan’ package (No-U-Turn-Sampler (NUTS)).

## Result

A summary of the COVID-19 confirmed cases by age group and symptom status onboard the Princess Cruises Ship is illustrated in Table 2. A total of 531 people had tested positive for the illness as of February 5, 2020. Out of 531 cases, three cases were aged 0-19 years, 117 were aged 20-58 years and 411 were aged 60 years and older. Finally, the crude asymptomatic ratio, a simple proportion of asymptomatic infections among all the infections was estimated as follows: 66.7% (95%Confidence Interval (95%CI): 9.4%, 99.1%) for aged 0-19 years, 30.8% (95%CI: 22.6%, 40.0%) for aged 20-58 years and 52.8% (95%CI: 47.9%, 57.5%) for aged 60 years and older (95%CI is based on binomial distribution).

**Table 1.** The whole voyage of the Princess Cruises Ship and important events related to the outbreak

| Day | Date | Event |
| --- | --- | --- |
| 1 | Jan/20/2020 | Departed from the Port of Yokohama<br>Index case embarked |
| 6 | Jan/25/2020 | Arrived at the Port of Hong Kong<br>Index case disembarked in Hong Kong<br>Departed from the Port of Hong Kong |
| 8 | Jan/27/2020 | Arrived at the Port of Chan May (Vietnam)<br>Departed from the Port of Chan May (Vietnam) |
| 9 | Jan/28/2020 | Arrived at the Port of Cai Lan (Vietnam)<br>Departed from the Port of Cai Lan (Vietnam) |
| 12 | Jan/31/2020 | Arrived at the Port of Keelung (Taiwan)<br>Departed from the Port of Keelung (Taiwan) |
| 13 | Feb/1/2020 | Arrived at the Port of Naha (Japan)<br>Departed from the Port of Naha (Japan)<br>Index case was confirmed |
| 16 | Feb/4/2020 | Arrived at the Port of Yokohama (Japan)<br>Passengers and crews were asked to stay on the ship for quarantine<br>Health status of all passengers and crew members were checked by questionnaire by quarantine officers |
| 17 | Feb/5/2020 | A lab-confirmed case of COVID-19 was detected.<br>Quarantine for 14 days begins at 7am.<br>Passengers requested to stay in their cabins. |
| 30 | Feb/18/2020 | End of quarantine |

**Table 2.** Summary of test positive COVID-19 cases by age group and symptom status onboard the Princess Cruises Ship.

| Age group | Symptomatic cases | Asymptomatic cases* | Total | Crude asymptomatic ratio ‡ | Persons aboard§ |
| --- | --- | --- | --- | --- | --- |
| 0-9 | 0 | 1 | 1 | 100%<br>(95% CI: 2.5%, 100%) | 16 |
| 10- | 1 | 1 | 2 | 50.0%<br>(95% CI: 1.3%, 98.7%) | 23 |
| 20- | 18 | 2 | 20 | 10.0%<br>(95% CI: 1.2, 31.7%) | 347 |
| 30- | 18 | 5 | 23 | 21.7%<br>(95% CI: 7.5%, 43.7%) | 429 |
| 40- | 18 | 7 | 25 | 28.0%<br>(95% CI: 12%, 49.4%) | 333 |
| 50- | 27 | 22 | 49 | 44.9%<br>(95% CI: 30.1%, 59.8%) | 398 |
| 60- | 73 | 56 | 129 | 43.4%<br>(95% CI: 2.5, 100%) | 924 |
| 70- | 92 | 136 | 228 | 59.6%<br>(95% CI: 53.0%, 66.1%) | 1015 |
| 80- | 27 | 25 | 52 | 48.1%<br>(95% CI: 34.0%, 62.3%) | 215 |
| 90- | 2 | 0 | 2 | 0%<br>(95% CI: 2.5%, 84.2%) | 11 |

§ As of February 5, 2020
‡ Proportion of asymptomatic cases among all the cases. CI: Confidence Interval (CI) is based on binomial distribution
\* Symptom status is based on the information at the time of specimen collection. There is a possibility that a fraction of asymptomatic cases develop symptom.

The observed and estimated daily number of cases among passengers and crew members from day 1 to day 29 (January 20, 2020–February 17, 2020) are shown in Figure 1. The total number of cases for all and by type (passengers and crew members) were estimated at 187.0 (95%CrI: 163.8, 212.3), 127.1 (95%CrI: 107.8, 148.9) and 59.6 (95%CrI: 47.3, 74.0), respectively. For comparison, the number of reported cases for all and by type were 197, 149 and 48 respectively. The corresponding percentage coverage of the 95%CrI of estimated data for observed data, the number of days where the model 95% CrI overlapped with the observed data, is 54% (15/28) for all, and 50% (14/28) and 43% (12/28) for passenger and crew, respectively.

**Figure 1.**
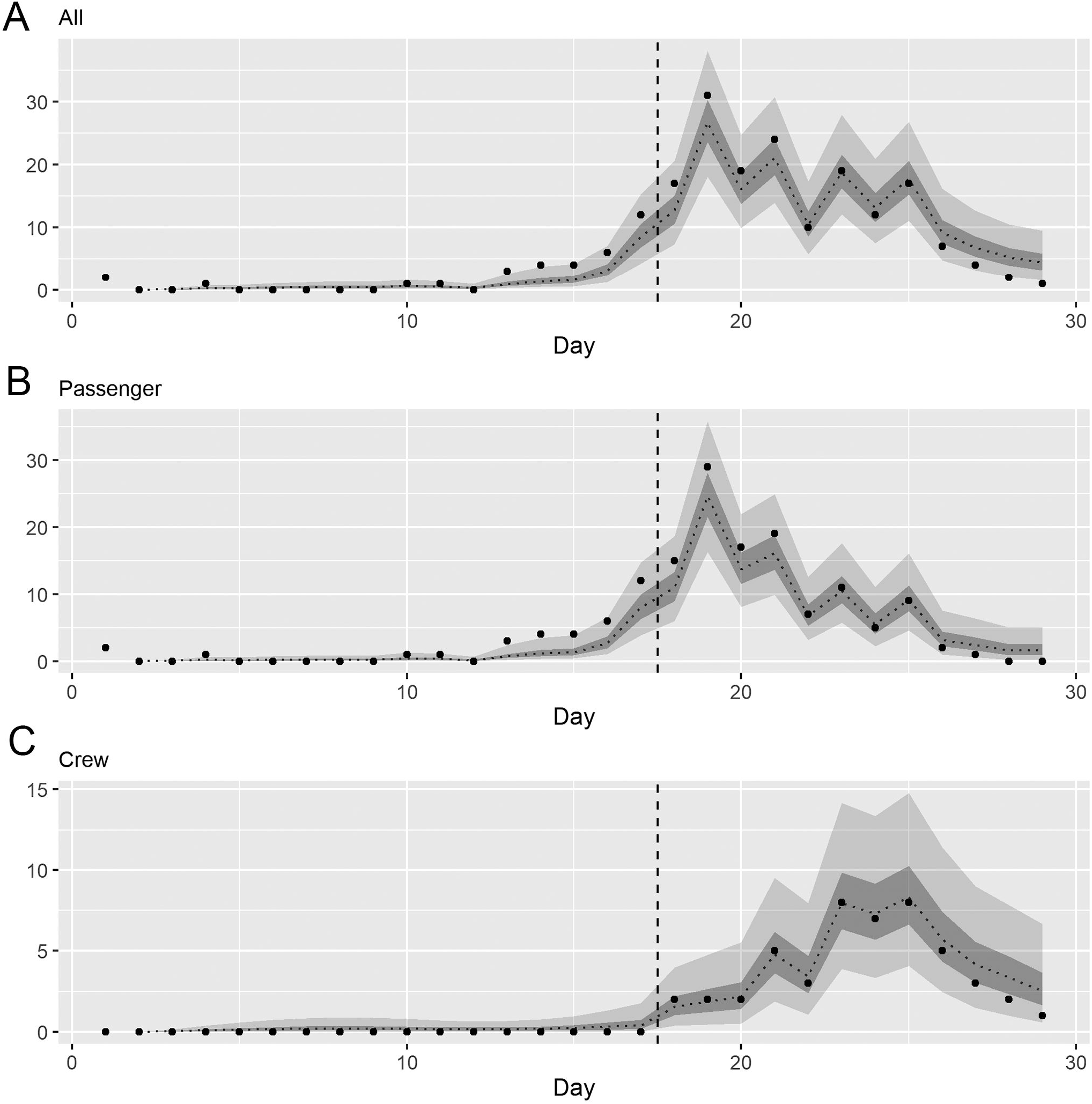
Observed and estimated number of the New Coronavirus (COVID-19) cases by type, onboard the Princess Cruises Ship, 2020 (n = 197) (A) - (C) Comparison of observed and predicted daily numbers of new cases by type. Black dots shows observed data, and light and dark indicates 95% and 50% credible intervals for posterior estimates, respectively. Day 1 on horizontal axis corresponds to January 20, 2020.

The time-dependent reproduction number for all and by type are presented in Figure 2. For all, *R*_t_ rapidly increased at around day 12 (January 31, 2020) and reached its maximum with the value of 11.2 (95%CrI: 7.5, 16.2) at day 19 (February 7, 2020). *R*_t_ for passengers presented a similar pattern with the value of 12.1 (95%CrI: 8.2, 17.2) at day 19 (February 7, 2020), while *R*_t_ for crew members only shows slight fluctuations with the largest value reaching 1.56 (95%CrI: 0.07, 7.55) at day 23 (February 11, 2020).

**Figure 2.**
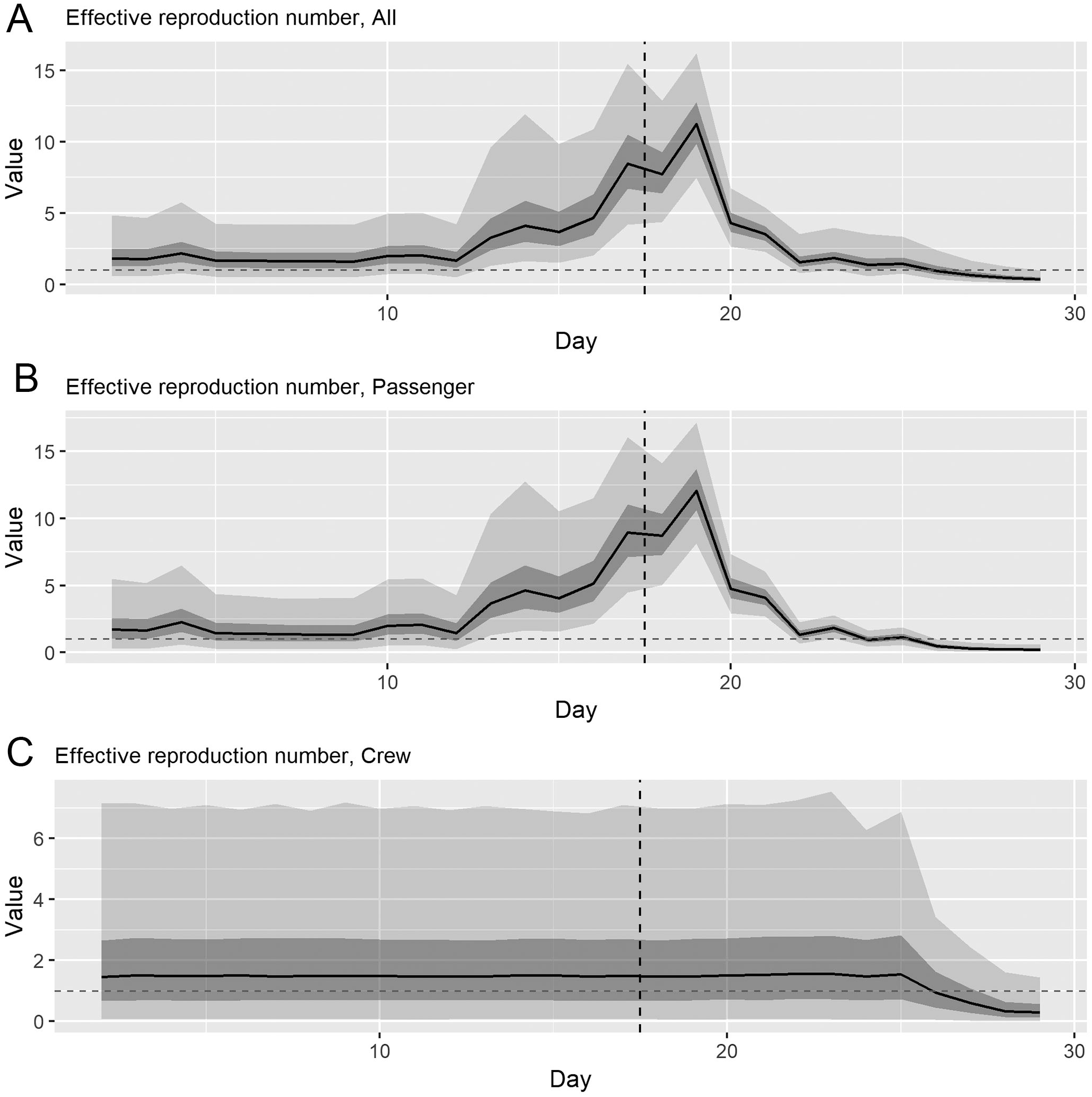
Time-dependent effective reproduction number of COVID-19 onboard the Princess Cruises Ship, 2020. The overall effective reproduction number were calculated from the dominant eigenvalue of next-generation matrix. Light and dark indicates 95% and 50% credible intervals for posterior estimates, respectively. Day 1 on horizontal axis corresponds to January 20, 2020. Horizontal grey dashed line shows the reproduction number at 1.0 for reference, below which incidence declines. Vertical dashed line indicates the day when quarantine was implemented (February 5, 2020).

Distributions of median *R*_t_ for entire study period for overall, for type and by each transmission type are shown in Figure 3. Median *R*_t_ are 5.8 (95%CrI: 0.6-11.0), 6.1 (95%CrI: 0.5, 11.8), 0.9 (95%CrI: 0.3, 1.5) for overall and by type (passenger, crew). Examining inter-type and within-type transmission, *R*_t_ estimates greatly vary across transmission types: 5.6 (95%CrI: 0.3, 10.9) for passenger to passenger, 0.6 (95%CrI: 0.1, 1.1) for passenger to crew, 0.5 (95%CrI: 0.2, 0.8) for crew to passenger, 0.5 (95%CrI: 0.3, 0.8) for crew to crew. Although vaccines are still in early development stages as of February 2020, based on our findings, the corresponding target vaccination coverage to contain the outbreak in this confined setting were estimated at 91% and 94% from the maximum value of the 50 percentile distribution and the 97.5 percentile distribution, respectively.

**Figure 3.**
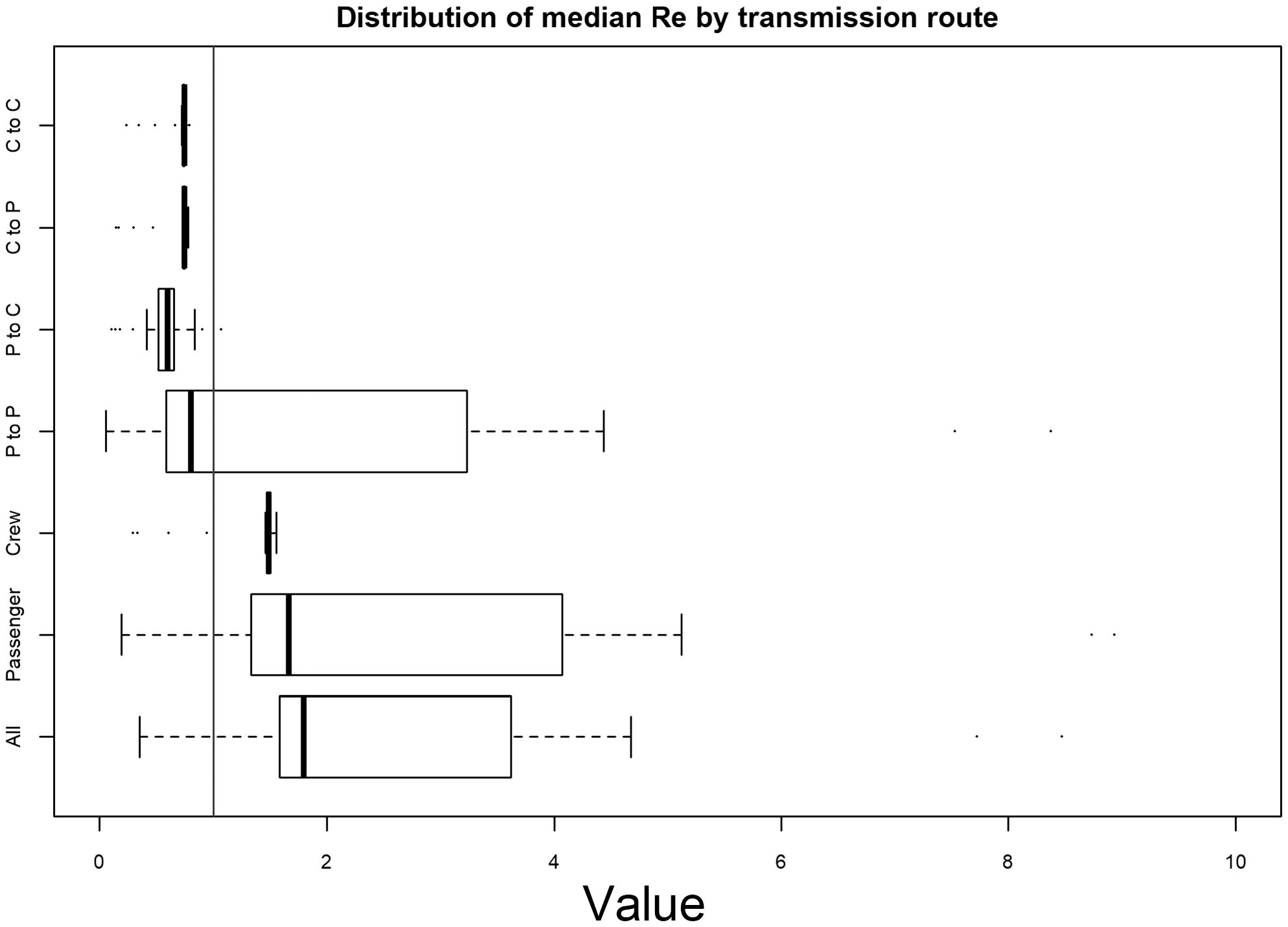
Distribution of median effective reproduction number for overall and by transmission type, onboard the Princess Cruises Ship, 2020. P: Passenger, C: Crew

Our latest estimate of the overall *R*_t_ is 0.35 (95%CrI: 0.02, 2.19), with only 2% of *R*_t_ estimates lying above the epidemic threshold of 1.0. Passenger and crew also have total (within-type and inter-type) *R*_t_ values largely below the epidemic threshold, with only small percentages at 0% and 9% above the epidemic threshold, respectively (Table 3).

**Table 3.** The latest estimate of median effective reproduction number and fraction of the density of *R* above the threshold of 1.0, February 18, 2020.

|  |  | Range | Percentage over 1.0 |
| --- | --- | --- | --- |
| All |  | 0.35 (95%CrI<sup>§</sup>:0.02, 2.19) | 2% |
| Passenger | Total <sup>‡</sup> | 0.19 (95%CrI: 0.00, 1.26) | 0% |
|  | Within<sup>¶</sup> (Passenger to Passenger) | 0.06 (95%CrI: 0.00, 0.73) | 0% |
|  | Inter (Passenger to Crew) | 0.10 (95%CrI: 0.00, 1.04) | 0% |
| Crew | Total | 0.30 (95%CrI: 0.00, 3.84) | 9% |
|  | Within (Crew to Crew) | 0.15 (95%CrI: 0.00, 1.92) | 1% |
|  | Inter (Crew to Passenger) | 0.24 (95%CrI: 0.00, 2.17) | 2% |

<sup>§</sup>CrI: 95% credibility intervals (CrI)
<sup>‡</sup>Total transmission includes within-type and inter-type transmission

## Discussion

This is the first study to assess the transmission potential of the COVID-19 outbreak that unfolded aboard the Diamond Princess Ship, January-February 2020. The overall mean reproduction number in the confined setting reached values as high as ∼11, which is higher than mean estimates reported from community-level transmission dynamics in China and Singapore in the range 1.1-7 [10-16]. However, following the implementation of the quarantine period, the overall *R*_t_ decreased substantially compared to values estimated during the early stage, but it exhibited fluctuations around the epidemic threshold, which likely prolonged the outbreak.

Our results indicate that *R*_t_ declined following the quarantine measures implemented by the Japanese government on February 5 relative to values during the early phase of the outbreak [3]. Importantly, only those passengers and crew who tested positive for the novel coronavirus were permitted to disembark the Diamond Princess Ship, with more than 80 percentage of the passengers and crews still on board as of February 18, 2020. Our latest overall estimate of *R*_t_ at 0.35 (95%CrI: 0.02, 2.19) with only 2% of *R*_t_ estimates lying above the epidemic threshold of 1.0 suggested a low probability of observing a subsequent outbreak aboard the Diamond Princess Ship.

Our findings indicate that the passenger-to-passenger transmission type dominated the transmission dynamics aboard the Diamond Princess Ship. The rapid increase in the overall *R*_t_ from day 12 (January 31, 2020) to day 19 (February 7, 2020) is greatly influenced by the increase in passenger-to-passenger transmission, and this time period covers the time lag between February 1, 2020 when the index COVID-19 case was reported and February 5, 2020 when the Japanese government requested the passengers to stay inside their cabin after the detection of a cluster of COVID-19 positive cases [3, 17]. Upon the implementation of the quarantine orders, the overall *R*_t_ and *R*_t_ for passengers aboard the Diamond Princess Ship declined thereafter, while *R*_t_ among crew maintained a steady level and only started to decline on day 25 of the outbreak (February 13, 2020). This is consistent with the fact that passengers staying inside their cabins led to a substantial decline in passenger-to-passenger transmission type except for their interaction with cabin mates. Indeed, the proportion of secondary infections inside the cabins increased from 7% (1/15) on February 6, 43% (3/7) on February 10, 2020 to 100% (1/1) on February 14, 2020 [3]. Thus, a high proportion of the symptomatic cases after the quarantine gradually shifted from largely passenger cases to crew cases. Indeed, despite their potential risk of acquiring the infection, crews had to continue to work to deliver services to isolated passengers. The distribution of the median Re by transmission type suggests that other transmission types likely contributed to this outbreak to some extent (Figure 3). Because one quarantine officer, one nurse, and one administrative officer contracted COVID-19 aboard the Diamond Princess Ship, the infection risk continued to be significant inside the Diamond Princess Ship.

As for the crude asymptomatic ratios by age groups, they show significant differences across age groups. However, these crude ratios are severely influenced by the timing of specimen collection relative to the infection time [19]. Hence, these ratios could be better ascertained if additional data with the timing of specimen collection becomes available.

Several limitations should be listed. First, a total of 103 laboratory-confirmed symptomatic cases with unknown onset dates were not incorporated in our analysis. Although the high proportion of unavailable data (34% (103/300)) likely influenced a downward bias in estimates of transmission potential, our estimates still point to the high transmission potential of COVID-19 inside confined settings. Second, it is possible that reporting delays could have influenced our latest estimates of the effective reproduction number. In fact, the US government recently sent a chartered flight to transport the American passengers on board the Diamond Princess Ship, and after the disembarkation process, a total of 14 American passengers tested positive for the disease [18] on February 16, 2020. This event also contributes to a downward bias in R although the number of cases does not exceed the peak of the outbreak, further supporting the potent transmissibility of COVID-19 in confined settings.

Our most recent estimate of the effective reproduction number of the ongoing COVID-19 epidemic on board the Diamond Princess Ship was largely below the epidemic threshold of 1.0, which suggested a very low probability of observing secondary outbreaks of the disease in the Diamond Princess Ship.

## Data Availability

The data that support the findings of this study are openly available in public sources.

## Notes

### Competing Interest Statement

The authors have declared no competing interest.

### Funding Statement

KM acknowledges support from the Japan Society for the Promotion of Science (JSPS) KAKENHI Grant Number 18K17368 and from the Leading Initiative for Excellent Young Researchers from the Ministry of Education, Culture, Sport, Science & Technology of Japan. GC acknowledges support from NSF grant 1414374 as part of the joint NSF-NIH-USDA Ecology and Evolution of Infectious Diseases program

